# Association between SARS-CoV-2 and Stroke: Perspectives from a metaumbrella-review

**DOI:** 10.1101/2024.10.01.24314742

**Authors:** Andreza Maria Luzia Baldo de Souza, Enoque Fernandes de Araújo, Nelson Carvas, Augusto César Raimundo, Antonio Carlos Pereira, Marcelo de Castro Meneghim

## Abstract

In the face of the global COVID-19 pandemic, the need arose to investigate potential complications associated with SARS-CoV-2, including the risk of Stroke.

**Objective:** This study aimed to verify the association between SARS-CoV-2 and the risk of Stroke, based on systematic reviews and meta-analyses, in order to assess the inclusion of the virus as a new risk factor for cerebrovascular diseases.

**Methods:** A metaumbrella was conducted, which included 34 systematic reviews, of which 4 were selected for the final analysis based on methodological quality and consistency. The analysis aggregated the results of 70 primary studies, considering different stroke subtypes and outcomes associated with COVID-19. Study heterogeneity was assessed using the I² index, and significance bias was verified using Egger’s test.

**Results:** The analysis showed that the severity of COVID-19 is significantly associated with an increased risk of stroke (eOR = 2.48; 95%CI: 1.55 – 3.95), particularly for ischemic stroke (eOR = 1.76; 95%CI: 1.11 – 2.80) and hemorrhagic stroke (eOR = 3.86; 95%CI: 1.79 – 8.33). Additionally, patients with cerebrovascular comorbidities had higher mortality (eOR = 2.48; 95%CI: 2.48 – 19.63), as did those who had previously suffered a stroke (eOR = 6.08; 95%CI: 3.73 – 9.91).

**Conclusion:** The association between SARS-CoV-2 and stroke was consistent and significant, suggesting that COVID-19 should be considered a new risk factor for cerebrovascular diseases. However, the high heterogeneity among the studies analyzed reinforces the need for further research to consolidate this relationship.

## Introduction

Responsible for millions of deaths annually, stroke is a global public health challenge^1-3^. It is a sudden neurological deficit, which can be transient or permanent, caused by a vascular injury that results in ischemia or hemorrhage in areas of the brain^2^. Stroke is a multifactorial disease, caused by a combination of modifiable, non-modifiable, and environmental risk factors ^1,4,5^.

The COVID-19 pandemic, caused by the SARS-CoV-2 virus, triggered a global health crisis^6,7^. Although it is primarily recognized for causing respiratory infections, recent studies have associated COVID-19 with increased risk of stroke^8,9,10^.

This association raises concerns about the mechanisms by which SARS-CoV-2 may be linked to neurological damage. Hypotheses include systemic inflammation, direct invasion of the nervous system by the virus, and complications of the immune response^12,13^. In addition, individuals with preexisting risk factors for stroke, such as hypertension and diabetes mellitus, seem to be more likely to develop more severe cases of COVID-19 and, consequently, a higher risk of stroke^14,15,16,17,18,19^.

This study aims to verify the association between SARS-CoV-2 and stroke, using systematic reviews as a guiding reference. The investigation seeks to contribute to the scientific debate on the possible inclusion of the virus as a risk factor for cerebrovascular diseases.

### Methodology

This study is characterized as an Umbrella Review^20^, which aims to synthesize the evidence from multiple systematic reviews^21,22^. The methodology used followed the PRIO-harms^23^ checklist to ensure the rigor and quality of the analysis. The formulation of the research question considered the following elements: population, phenomenon of interest, result, context, type of overview and general objective^24,25,26^. Based on the hypothesis that SARS-CoV-2 infection is associated with increased risk of stroke, the following guiding question was formulated: "Does the association between SARS-CoV-2 and stroke presuppose the need to include it as a new risk factor in the list for cerebrovascular disease?". The protocol of this study was registered in the International Prospective Register of Systematic Reviews, under number CRD42022323750.

### Search Strategy

Studies published in English, Spanish, or Portuguese, from March 2020 to March 2023, that address the association between COVID-19 and ischemic or hemorrhagic stroke, small or large vessels, in any age group, were selected. The databases consulted were PubMed/MEDLINE, LILACS, Scopus, and Web of Science. The search strategy used a strategic combination of terms and keywords in all three languages. The terms used were: "Stroke", "COVID-19", "Neurological Complications", "Systematic review"; "Cerebrovascular Accident", "COVID-19", "Neurological Complications", "Systematic Review"; "Stroke", "COVID-19", "Neurological Complications", "Systematic Review"

To complement and broaden the search, the following terms were used in different combinations, using Boolean operators to improve the results: (STROKE* OR CEREBROVASCULAR* OR NEUROLOGICAL*) AND (COVID* OR SARS-CoV-2*) AND (SYSTEMATIC* AND REVIEW*); (("Stroke" OR "Stroke") AND ("systematic review" OR "systematic review" OR "systematic review")) AND ("SARS-CoV-2").

### Selection criteria

Scientific articles were selected that include systematic reviews, systematic reviews with meta-analysis of case studies, case series, case-control studies and, preferably, randomized and prospective and retrospective cohort studies. Reviews that were not available in full, incomplete manuscripts, studies outside the context of systematic review, and non-original research articles, such as editorial comments, opinion articles, letters, protocols, reports, and book chapters, were excluded. Also excluded were reported non-clinical features, such as non-neurological complications, as well as studies that presented a diagnosis of COVID-19 without any reports of stroke as a complication.

### Data extraction

The selection of articles was carried out by two independent reviewers (AMLBS and EFA) in two stages. First, the titles and abstracts were independently evaluated, and any disagreements were resolved by consensus. Then, the full text of the selected articles was analyzed in the same way, with consensus being used to resolve disagreements.

The agreement between the reviewers was assessed using Cohen’s Kappa coefficient^27^. In the screening phase of titles and abstracts, the Kappa coefficient was 0.62511, indicating a substantial agreement among the reviewers. This result suggests that the selection criteria were well defined and understood, resulting in a consistent initial selection of studies.

The use of the Covidence^28^ software brought significant benefits to the review process, facilitating the organization and analysis of the data, including the calculation of the Kappa index and the generation of the PRISMA flowchart. This online tool allowed for real-time collaboration between reviewers, simplifying the resolution of disagreements and ensuring the transparency of the process.

### Quality assessment

The methodological quality of systematic reviews was assessed using the ROBIS^29^ tool, a validated and widely used instrument to assess the risk of bias in systematic reviews in healthcare. The ROBIS tool is especially useful for evaluating reviews that address interventions, diagnosis, prognosis, and etiology, and is therefore suitable for the scope of this study.

The evaluation process with the ROBIS tool is divided into three main phases: Phase 1: Assessment of the relevance of the systematic review to the research question. In this step, it is verified whether the selected systematic review directly addresses the research question of the Umbrella Review.

Phase 2: Identification of concerns with the systematic review process. This phase investigates four critical domains that may be sources of bias: Study eligibility criteria: Evaluates whether the inclusion and exclusion criteria of the primary studies were adequate and well-defined. Identification and selection of studies: Analyzes the search and selection process of studies, checking whether there was a risk of publication bias. Data collection and study evaluation: Examines the quality of data collection and the assessment of risk of bias in primary studies. Synthesis and findings: Evaluates the presentation and synthesis of the results, considering the heterogeneity between the studies.

Phase 3: Judging the overall risk of bias for the systematic review. Based on the analyses of the previous phases, the overall risk of bias of the systematic review is classified as low, high, or unclear.

### Data analysis

Initially, for each identified factor, being evaluated in more than one individual study, we performed a separate random-effects meta-analysis to obtain a pooled estimate of the effect size, which we assumed would follow a normal distribution with variance equal to the sum of the weights of the studies^30^ (method of DerSimonian and Laird, 1986). The results of the meta-analyses were the effect sizes with their corresponding 95% confidence intervals (95% CI) and p-values, as well as the statistics needed to assess the level of evidence. We used the effect size measure used in each original meta-analysis (i.e., RR, OR, or SMD) and calculated the OR equivalents (eOR) for all effect size statistics.

We evaluated the heterogeneity between studies with the *I^2^ index* ^31^. I2 values > 50% indicated great heterogeneity ^33^. We also assessed whether there was evidence of effects from small studies using the Egger test ^33^, where statistical significance would mean potential publication bias ^34^.

In addition, a rating system for the strength of evidence was used, which has been widely used in previous umbrella reviews^35,36^. Specifically, we classified the levels of evidence of the significant associations between each factor into convincing evidence (class I), highly suggestive (class II), suggestive (class III), or weak evidence (class IV). Convincing evidence would require a number ≥ 10 studies, a number of cases ≥ 500, I2 ≤ 50%, and no signs of influence of small studies in the meta-analysis (Egger test ≥ 0.10). The suggestive evidence required a number ≥ 10 studies, a number ≥ 400 cases, an Egger test with a p-value ≥ 0.10, and I2 ≤ 50%. Weak evidence with a case count ≥ 300, Egger’s test with a P-value ≥ 0.10, I2 ≤ 75%, and very weak evidence did not require a specific number of cases and p<0.05.

Finally, the meta-analyses were repeated estimating heterogeneity with the Hartung-Knapp-Sidik-Johkman method for random effects. This method estimates variance as the weighted mean square error divided by degrees freedom and assumes a distribution t ^37,38,39^. The main difference between a normal distribution and a distribution t is that in the former, we assume that we can know variance, while in the latter, we do not make this assumption, as indeed is the case. This difference can be negligible when the number of studies is large, but it can be relevant when the number of studies is small. All analyses were performed with version 1.0.11 of the metaumbrella package, implemented in R environment.

## Results

### Identification and Selection of Studies

From an initial search in databases and registries, 2,490 studies relevant to the investigation of the association between COVID-19 and stroke were identified. After removing 1,289 duplicate references, 1,201 studies went through the screening process. Of these, 141 were excluded because they did not meet the relevance criteria, focusing mainly on management or medications, which was not the focus of this study. This resulted in the detailed evaluation of 1,060 studies for their eligibility.

Of these 1,060 studies, 1,026 were excluded for various reasons, including focusing on non-neurological manifestations of COVID-19, specific non-pertinent populations, medical conditions unrelated to COVID-19, inadequate methodologies, or unrelated interventions.

At the end of this process, 34 studies were considered eligible. Of these, four studies were selected for analysis in the metaumbrella, based on high methodological quality and consistency with the established criteria (Figure 1).

**Figure 1.**
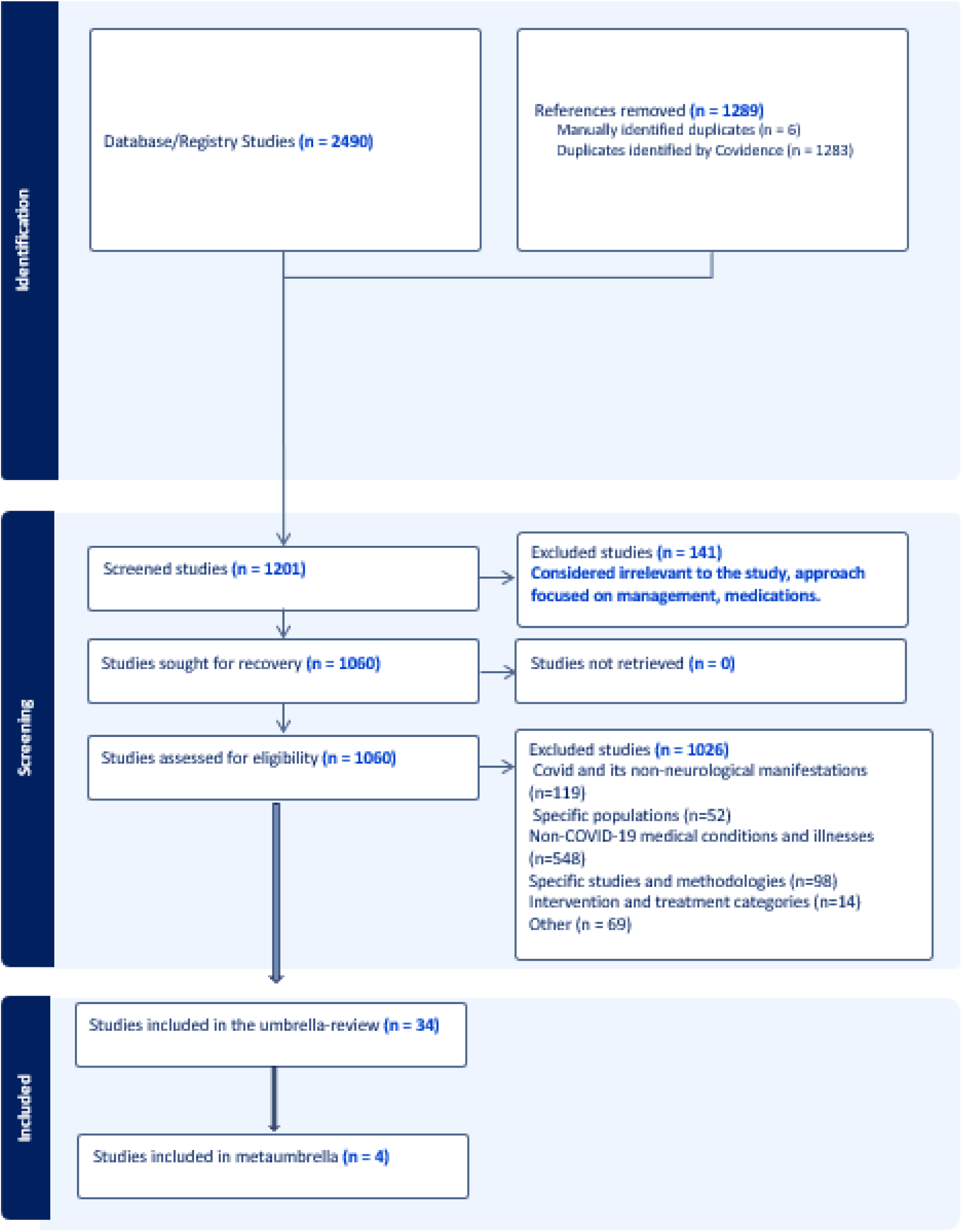
Prism.

### Characteristics of the Included Studies

The main characteristics of the 34 studies initially found demonstrate an important cohesion in the demographic and geographic profiles of the patients evaluated. The mean age of the patients was 61.2 years, which indicates that the study population consisted predominantly of individuals in an age group at higher risk for stroke. In addition, there was a clear predominance of males, with an average of 59.9% of participants being men. This disparity may be associated with men’s greater susceptibility to developing severe forms of COVID-19 and its complications, including stroke.

Geographically, the studies were conducted in a variety of countries, reflecting the global spread of the pandemic. Among the most frequently cited places are the United States, Italy, India, Brazil, and Spain, with particular emphasis on China. This country has emerged as the most frequently represented location, possibly due to the initial and significant impact of the COVID-19 pandemic on its territory, which has led to increased production of data and studies on the neurological complications associated with SARS-CoV-2.

### Risk of Bias Assessment

Figure 2 shows the evaluation of the methodological quality of the 34 studies included in the umbrela review, using the ROBIS tool. Most studies were at low risk of bias in criteria such as eligibility, identification and selection of studies, and data collection. However, some studies have shown uncertain or high risks, particularly in the selection of studies and the synthesis of results.

**Figure 2.**
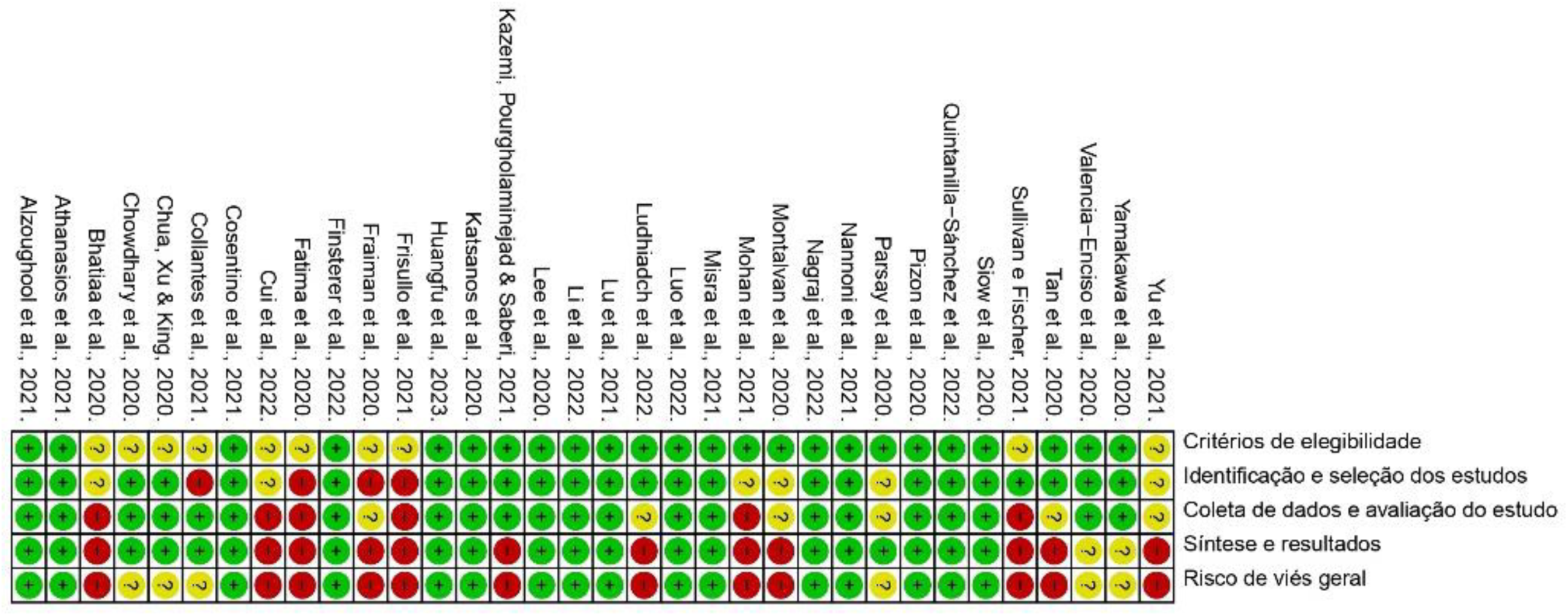
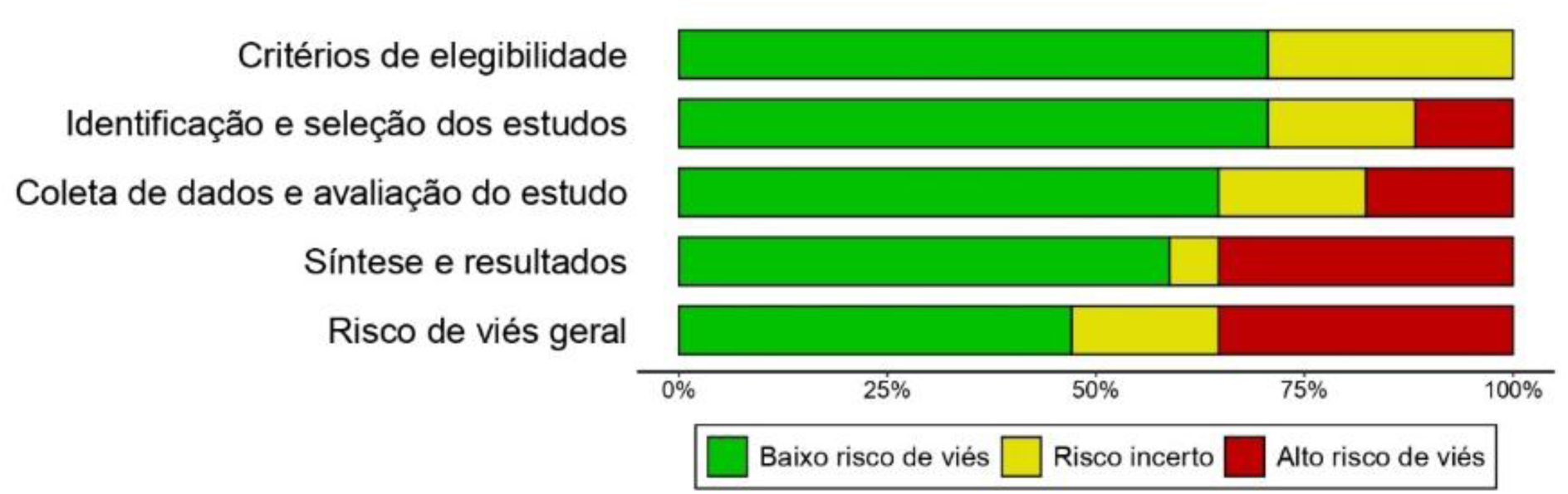
Quality of the ROBIS studies.

Among the four studies selected for the meta-umbrella, the assessment of bias was predominantly favorable, with all being classified as low risk in terms of overall bias.

### Metaumbrella Results

The results of the metaumbrella (Figure 3), which included four systematic reviews with meta-analysis, covered a total of 70 primary studies that evaluated the association between COVID-19 and stroke in five different study subjects. These objects of study were:

1. "COVID-19 severity and stroke risk": The meta-analysis showed that there is a significant association between COVID-19 severity and increased stroke risk, with an odds ratio (eOR) of 2.48 (95% CI: 1.55 – 3.95). This indicates that patients with severe COVID-19 are significantly more likely to develop stroke compared to those with less severe forms of the disease.
2. "COVID-19 and ischemic stroke risk": A significant association was found between COVID-19 and a higher risk of ischemic stroke, with an eOR of 1.76 (95% CI: 1.11 – 2.80). This suggests that COVID-19 infection may be a risk factor for developing ischemic stroke.
3. "COVID-19 and hemorrhagic stroke risk": The analysis also revealed an association between COVID-19 and increased risk of hemorrhagic stroke, with an eOR of 3.86 (95% CI: 1.79 – 8.33). This finding indicates that, in addition to ischemic stroke, COVID-19 may also be related to an increased risk of hemorrhagic stroke.
4. "Cerebrovascular comorbidity and mortality in patients with COVID-19": Patients with cerebrovascular comorbidity who contracted COVID-19 had a higher mortality compared to those who did not have a stroke, with an eOR of 2.48 (95% CI: 2.48 – 19.63). This result highlights the adverse impact of pre-existing cerebrovascular conditions on the survival of COVID-19 patients.
5. "COVID-19 and stroke mortality": Mortality was significantly higher among COVID-19 patients who already had a history of stroke, with an eOR of 6.08 (95% CI: 3.73 – 9.91). This data underlines the severity of the impact of COVID-19 on patients who had already suffered a stroke before.

**Figure 3.**
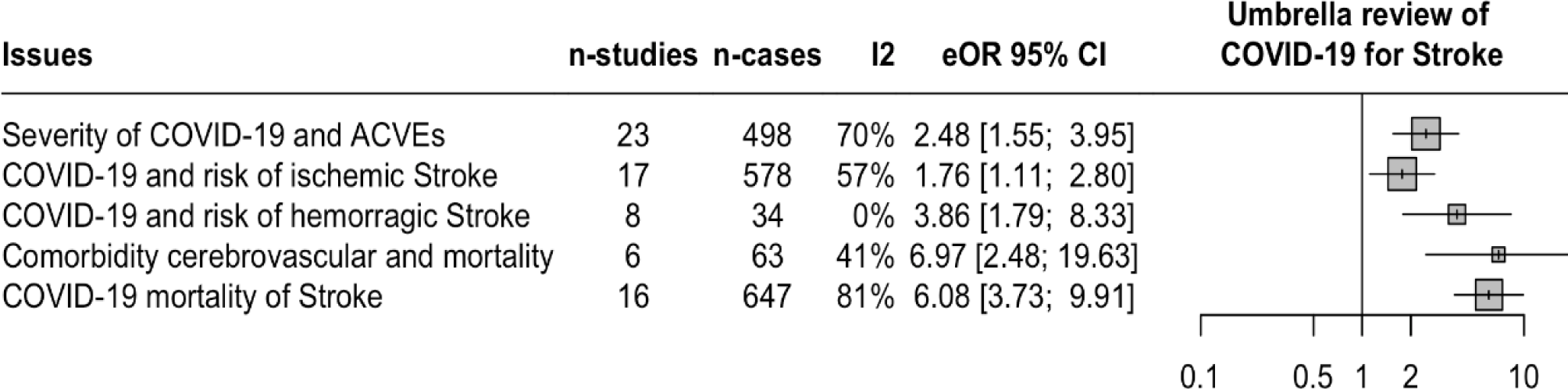
Meta Umbrella showing the association between COVID-19 and stroke.

In addition to these results, an overlap of two primary studies (Qureshi^40^ and Merkler^41^) was observed (Figure 4) in three distinct systematic reviews (Cui 2022^42^, Huangfu 2023^43^, and Quintanilla-Sánchez 2022)^44^. The overlap of these studies in the different reviews indicates that they are important and frequently cited references in the literature on the relationship between COVID-19 and stroke. These findings reinforce the strong association between COVID-19 infection and the risk of different types of strokes, as well as highlight the higher mortality associated with stroke in patients with COVID-19.

**Figure 4.**
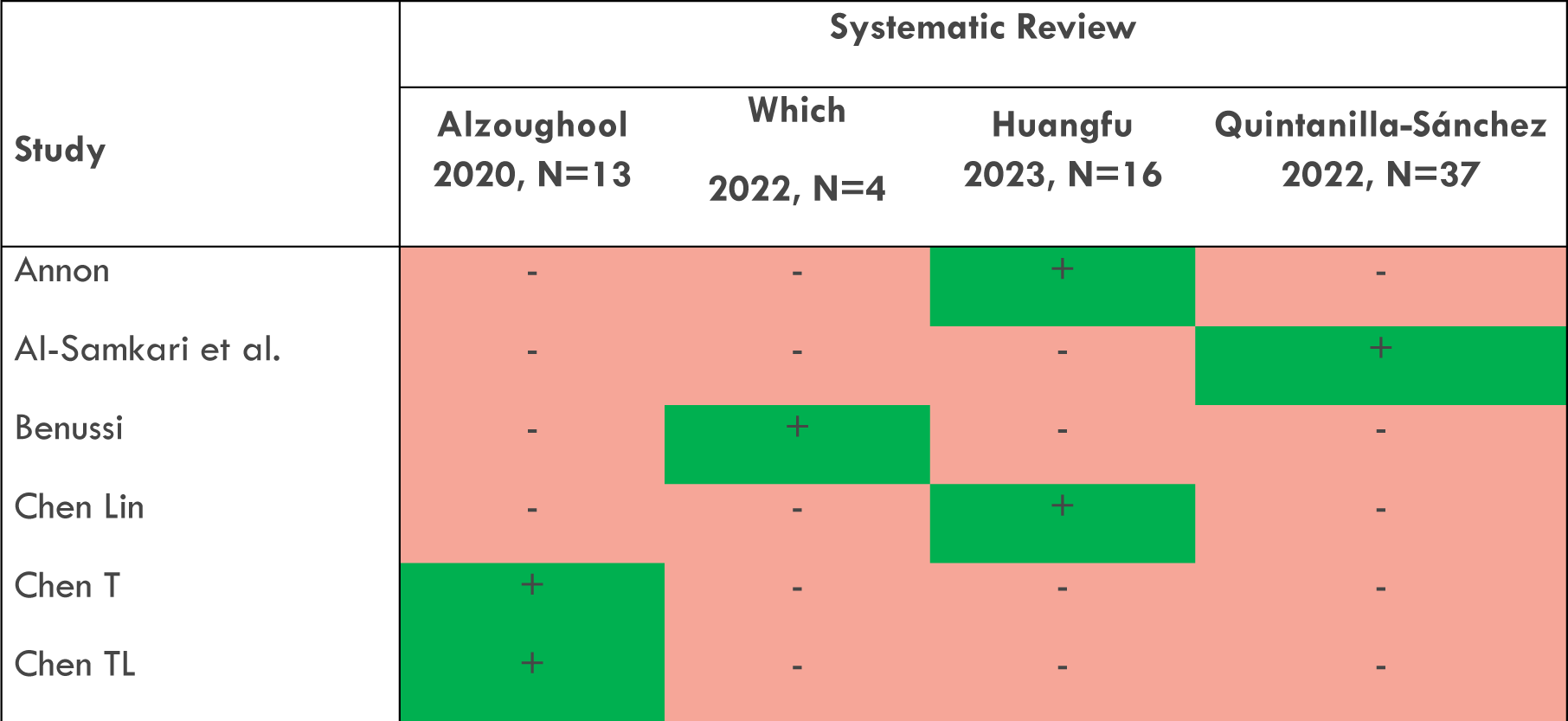
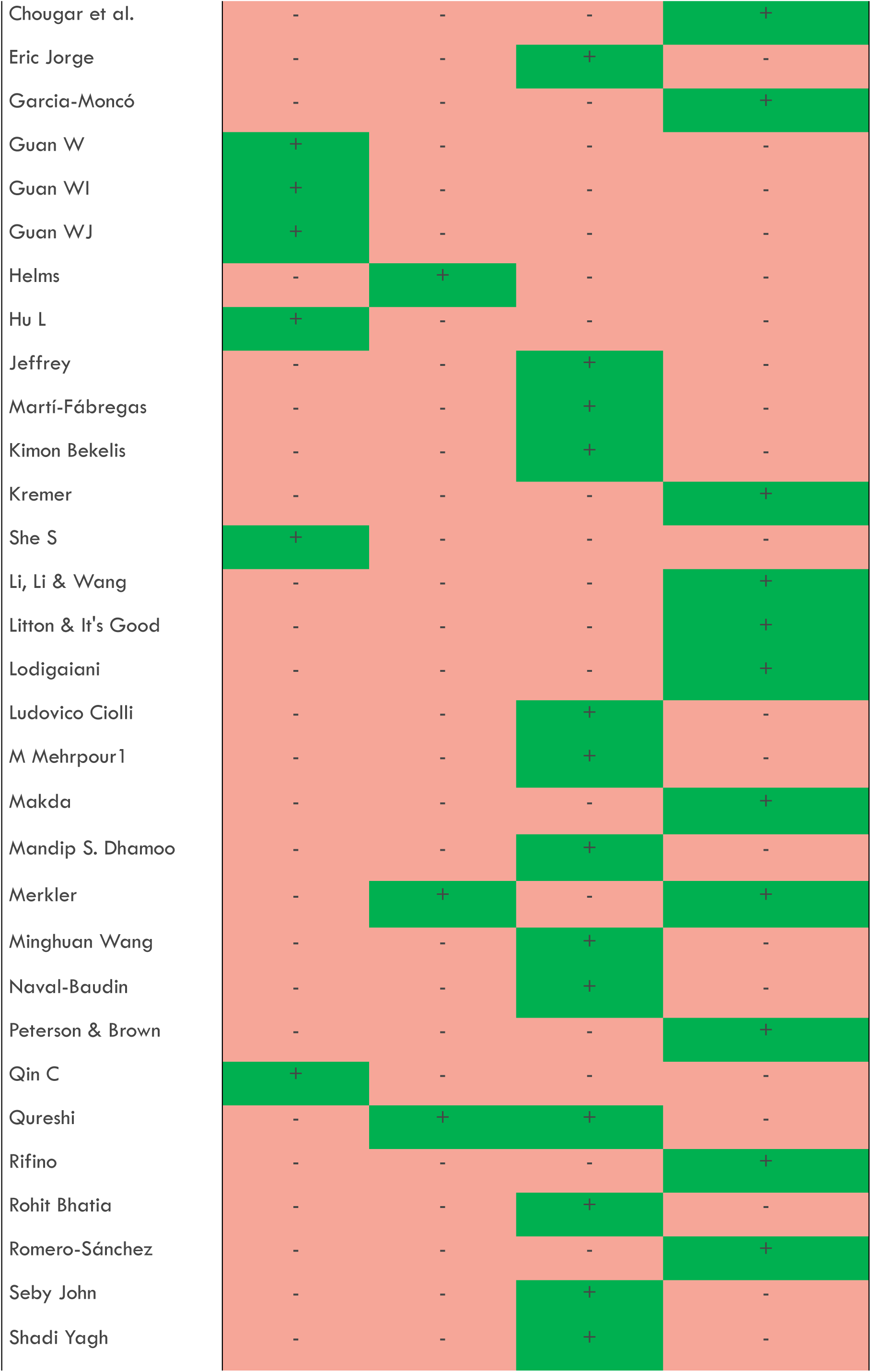
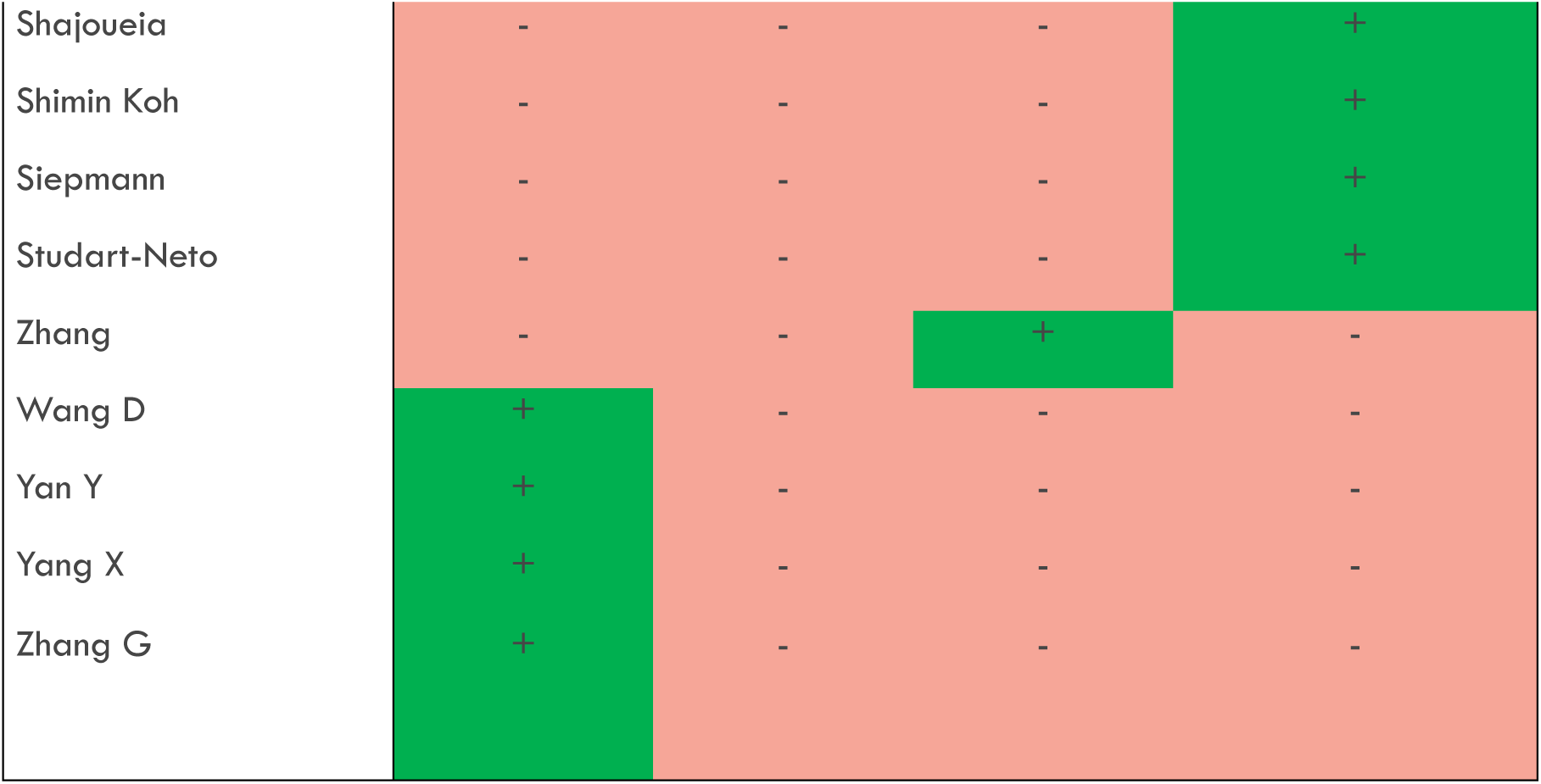
Matrix of overlapping studies in the systematic review.

### Analysis of heterogeneity and bias

Table 1, which presents the metaumbrella stratified by the classification of the evidence, it was observed that two of the study objects showed low heterogeneity, with *I² values* below 50%. This indicates that the variability between the studies included in these study objects was relatively low, suggesting a greater consistency in the results. In particular, the "Severity of COVID-19 and stroke" and "Cerebrovascular comorbidities and mortality" demonstrated this characteristic of low heterogeneity, which strengthens confidence in the interpretation of the observed effects.

**Table 1.** Total participants and P values from the Egger and JK test.

| Object of study | Number of patients | Number of Cases | Number of Controls | Egger p | ESB p |
| --- | --- | --- | --- | --- | --- |
| COVID-19 Severity and LVCAs | 15279 | 498 | 14781 | 8.45E-01 | 5.14E-01 |
| COVID-19 and ischemic stroke risk | 36154 | 578 | 35576 | 2.84e-01 | 1.59e-02 |
| COVID-19 and hemorrhagic stroke risk | 1303 | 34 | 1269 | 1.81e-01 | 7.31e-01 |
| Cerebrovascular comorbidities and mortality | 2271 | 63 | 2208 | 8.07e-01 | 9.56e-01 |
| COVID-19 and stroke mortality | 4781 | 647 | 4134 | 1.10e-01 | 0.252e-02 |

Egger p = Egger's test for bias due to the influence of small studies
ESB p = test for bias due to Statistical Excess Significance.

None of the study subjects analyzed showed the effect of small studies, as indicated by the non-significant values of the Egger test (Egger p). This suggests that the results of the meta-analyses were not significantly influenced by smaller studies, which could skew the conclusions.

However, two study subjects showed excess significance bias (ESB), which was identified by significant p-values: "COVID-19 and stroke mortality" (p = 0.0252) and "COVID-19 and risk of ischemic stroke" (p = 0.0159). This bias occurs when there is an excessive number of studies with positive results relative to what would be expected by the normal distribution of true effects, indicating that findings in these domains should be interpreted with caution.

The five study objects evaluated had a statistically significant effect size (p<0.05), which reinforces the validity of the findings. However, based on the criteria previously established for the classification of evidence, three of these study objects were classified as having weak evidence. This reflects limitations such as possible biases or inconsistencies in the results, suggesting the need for further studies to confirm these associations.

Figure 5 complements this information by stratifying the metaumbrella by the classification of evidence, visually highlighting the relative robustness of each object of study. This detailed analysis allows for a more nuanced understanding of the effects of COVID-19 in relation to stroke, while identifying areas where the evidence is weaker and where future studies could be more informative.

**Figure 5.**
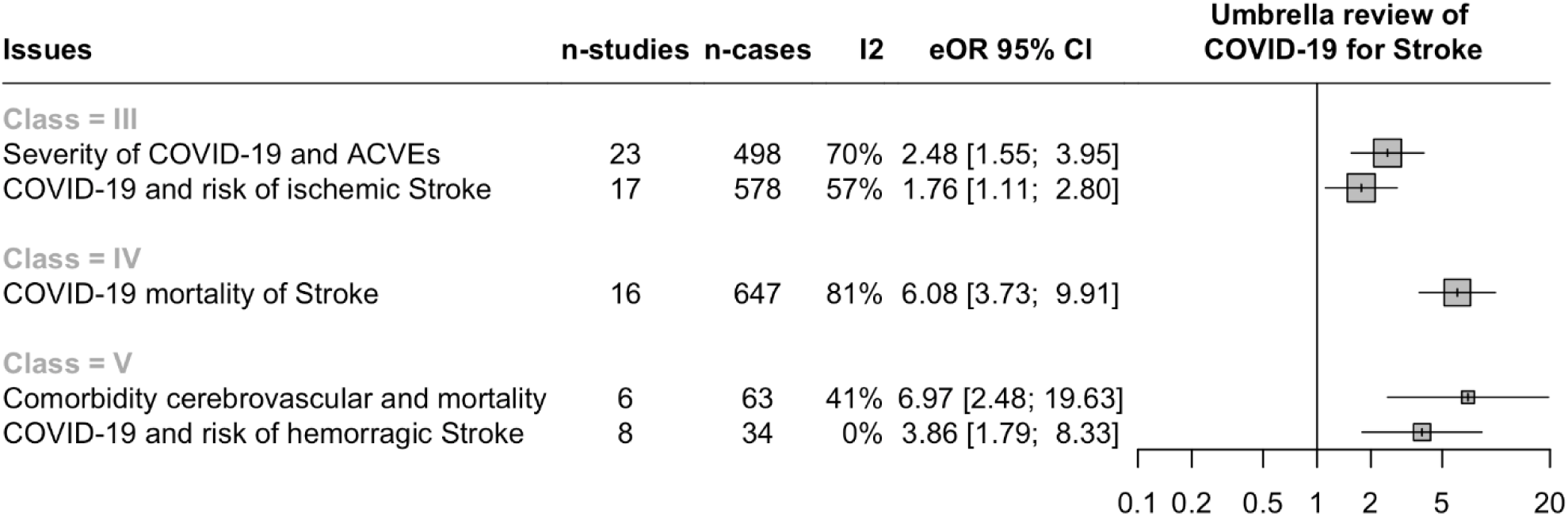
Umbrella Goal stratified by Evidence Classification.

## Discussion

This study started from the hypothesis that SARS-CoV-2 infection is associated with increased risk of stroke and sought to answer the guiding question: "Does the association between SARS-CoV-2 and stroke presuppose the need to include it as a new risk factor in the list for cerebrovascular disease?" since it proposes to deepen the understanding of the influence of COVID-19 on stroke risk, a global public health problem that is among the main causes of death and disability^45,46^.

A point to consider is the incidence of stroke in patients with COVID-19, which is significantly higher than in patients infected with other coronaviruses, suggesting a specific pathological mechanism associated with SARS-CoV-2 that predisposes to stroke^47^.

The meta-umbrella methodology used in this study offers significant advantages over individual systematic reviews. The comprehensive analysis of multiple meta-analyses, considering the overlap of primary studies, as exemplified by the inclusion of the study by Qureshi ^40^, Merkler^41^ in different analyses, ensures greater robustness and reliability of the results. The convergence of evidence from multiple sources, confirming the association between COVID-19 and different stroke subtypes, as well as associated mortality, strengthens the conclusion that COVID-19 represents an independent risk factor for stroke.

The finding of a link between COVID-19 and increased risk of stroke, especially the ischemic type, corroborates the literature that points to prothrombotic mechanisms induced by the virus^11,12,14^. Among these mechanisms, SARS-CoV-2 infection stands out, which triggers an acute inflammatory response that can result in endothelial dysfunction and a prothrombotic state^42^.

COVID-19 is associated with a state of hypercoagulability, increasing the risk of blood clots forming that can obstruct blood vessels in the brain, leading to stroke. SARS-CoV-2 can directly damage endothelial cells, which line blood vessels, making them more prone to the formation of these clots^48,49^. The high incidence of thrombotic complications in patients with severe COVID-19 reinforces the link between coagulation and viral infection, consolidating the relevance of the findings of this study.

The identification of SARS-CoV-2 as a risk factor for stroke has crucial implications for the prevention, diagnosis, and treatment of this condition^50^. It is essential to integrate this information into clinical practice, adopting measures such as: monitoring patients with COVID-19 for neurological symptoms, especially those at high risk of stroke, considering prophylactic anticoagulation in patients with COVID-19 and high risk of thromboembolic events, implementing screening protocols for stroke in patients hospitalized with COVID-19, especially in those with additional risk factors for cerebrovascular diseases^51^.

Some limitations should be considered, such as the heterogeneity in the diagnostic criteria for stroke among the studies, the variability in the sample size, and the inadequate control of confounding factors, such as hypertension and diabetes.

Future prospective, multicenter studies are essential to investigate the mechanisms underlying the association between COVID-19 and stroke in greater depth, to develop comprehensive clinical guidelines for the management of patients with COVID-19 and stroke risk, and to evaluate the efficacy of preventive interventions, such as anticoagulation, in reducing the incidence of stroke in patients with COVID-19.

## Conclusion

The association between SARS-CoV-2 and stroke was consistent and significant, suggesting that COVID-19 should be considered a new risk factor for cerebrovascular diseases. However, the high heterogeneity among the studies analyzed reinforces the need for further research to consolidate this relationship.

## Data Availability

All worksheets can be provided by the corresponding author through the email request

## Conflict of Interest Statement

The author(s) declared that there are no potential conflicts of interest with respect to the research, authorship and/or publication of this article.

## Financing

The author(s) did not receive financial support for the research, authorship and/or publication of this article.

All worksheets can be provided by the corresponding author through the email request

## Notes

### Competing Interest Statement

The authors have declared no competing interest.

